# Photoacoustic imaging in mitochondrial disease

**DOI:** 10.64898/2026.03.10.26347962

**Authors:** Thomas R. Else, Lorna Wright, Katherine Schon, May Yung Tiet, Chloe Seikus, Elizabeth Ashby, Charlotte Addy, Heather Biggs, Emma Harrison, Jelle van den Ameele, Patrick F. Chinnery, Sarah E. Bohndiek, Rita Horvath

## Abstract

Mitochondrial diseases are a diverse group of inherited neuromuscular disorders leading to progressive disability and early mortality. Mitochondrial myopathy is a common feature of mitochondrial disorders, affecting most patients. Assessment of disease progression and treatment efficacy in mitochondrial disease trials has often relied on muscle biopsies, however, these are increasingly considered unfavourable by patients. Imaging biomarkers of disease could reduce the patient burden, enabling non-invasive longitudinal monitoring of molecular information.

Photoacoustic imaging combines the molecular sensitivity of light absorption with the deep tissue imaging capabilities of ultrasound, enabling a safe and fast imaging technique. Tuning the wavelength of light allows for the detection of molecular constituents such as oxy- and deoxy-haemoglobin, lipids, and water. These signatures may reflect underlying pathophysiological alterations and serve as valuable indicators of disease state and progression.

We conducted an exploratory study of a photoacoustic imaging dataset in patients with mitochondrial myopathy due to the m.3243A>G mt-tRNALeu mutation and compared to healthy volunteers. We generated photoacoustic measurements at wavelengths in the near infrared, comparing absolute values and ratios derived in the bicep muscle. Confounding factors such as skin colour and sex were considered, and we ensured that these parameters were matched in healthy volunteers and patients. We identified significant differences between patients and controls, revealing changes in ratios between water and total haemoglobin, lipid and total haemoglobin, and lipid and water content. This study highlights the promise of photoacoustic imaging as a novel imaging biomarker in mitochondrial myopathies, paving the way for larger scale studies.

## Introduction

Mitochondrial diseases are a diverse group of inherited neuromuscular disorders that usually affect organs with high energy demand, including the skeletal muscle.^1^ While advances in genomics now allow molecular diagnosis in the majority of mitochondrial disease patients, treatments remain largely supportive, leading to progressive disability and early mortality.^2^ The development of new treatments is impeded by the scarcity of patients^2^ and limitations in the methods used to assess disease status. The rarity and variable clinical presentation of these disorders mean that small and heterogeneous patient cohorts are required in both natural history and interventional studies, which can make it challenging to draw definitive conclusions. Molecular biomarkers of mitochondrial diseases are typically derived from invasive and painful muscle biopsies, which reduces study participation and makes longitudinal studies more challenging.^3^

To enable faster and more frequent longitudinal patient monitoring, non-invasive imaging biomarkers have been proposed for evaluating mitochondrial function. Structural magnetic resonance imaging (MRI) can be used to assess the presence of a range of disease-associated pathologies in the brain and in skeletal muscles.^4,5^ Functional 31-P or 1-H magnetic resonance spectroscopy (MRS) in brain or skeletal muscle can be used to directly interrogate the concentration and spatial distribution of metabolites in patients with mitochondrial disorders.^4^ For example, these patients may display lower concentrations and different compartmentalisation of adenosine triphosphate, seen with 31-P MRS, or a buildup of lactate particularly in the brain and in muscles after exercise, seen with 1-H MRS.^6^ Alternatively, positron emission tomography (PET) can be used together with targeted radiotracers. For example, [11C]PK11195 targets the mitochondrial outer membrane translocator protein and is a general marker of glial activation, and signal changes in the brain have been shown to correlate with mitochondrial disease severity.^4,5,7,8^

Both MRI and PET, however, suffer from high costs and slow acquisition times, while PET additionally requires the use of ionising radiation. They are also challenging to perform in children, where mitochondrial diseases can be particularly troublesome, often requiring anaesthesia.^9^ Therefore, the use of a portable imaging modality instead, like ultrasound, may be preferable. Unfortunately, ultrasound does not provide specific molecular information and instead relies on structural measures such as muscle thickness or volume, yielding low specificity.^10^ Thus, there is an unmet need for an affordable, bedside imaging modality capable of providing molecular assessment of tissue composition to support longitudinal monitoring and facilitate streamlined evaluation of novel therapies.

We hypothesised that photoacoustic imaging (PAI) could offer a compelling alternative to existing radiological imaging modalities for mitochondrial disease assessment. By combining molecular sensitivity through light absorption with the deep tissue imaging capability of ultrasound, it enables a safe, non-invasive, contrast agent-free and relatively low-cost approach to tissue molecular imaging.^11^ It is making its way towards clinical translation, having recently obtained regulatory approval in the United States, the European Union, and Japan.^12^ Through its sensitivity to melanin, oxy- and deoxy-haemoglobin, lipids and water, photoacoustic imaging has shown promise across several applications, from cancer to neuromuscular disease and inflammation.^12–15^ Changes in tissue haemoglobin and lipids measured with PAI have been detected in patients with Duchenne muscular dystrophy,^14^ spinal muscular atrophy,^10,16,17^ and late-onset Pompe disease^18^ and a high degree of reproducibility has been demonstrated.^19^

Here, we present an exploratory analysis of PAI data acquired in skeletal muscle of patients with mitochondrial disease and compare the imaging features to those seen in healthy volunteers. While controlling for the potentially confounding factors of sex and skin colour,^20–24^ we show that even in a relatively small (n=11) number of patients, there are significant changes in the photoacoustic spectra at wavelengths associated with total haemoglobin, lipids and water, highlighting the potential for PAI to play a role as a biomarker in mitochondrial disease.

## Materials and methods

### Study design

The study was conducted between May 2021 and October 2025, following approval by local Research Ethics Committees (Ref: 19/EE/0157; Ref: 13/YH/0310; Ref: 23/EE/0019).^21^ Written informed consent was obtained from all study participants. The study was conducted in accordance with the Declaration of Helsinki. As skin colour is known to adversely affect PAI data quantification, we used a previously developed linear model^21^ to exclude all healthy volunteers outside the range of that observed in the m.3243A>G patients. We excluded data from healthy volunteers with a higher signal at 700 nm (strongly correlated with independent assessment of melanin concentration)^21^ in the skin surface region of interest than the maximum value in the m.3243A>G patients group. After ensuring that there was no significant difference between the epidermis photoacoustic signal between the cohorts, there were 11 patients with m.3243A>G-associated mitochondrial disease and 21 healthy volunteers included in the analysis presented here.

### Photoacoustic imaging protocol

Ultrasound and PAI data were acquired from the bicep muscle (Figure 1A) in all participants to examine a range of potential biomarkers (Figure 1B) that could be mapped using PAI (Figure 1C). Ultrasound images were used as a structural reference for drawing regions of interest (Figure 1C). Three different generations of a commercial PAI system, the iThera Acuity Echo (iThera Medical GmbH, Munich, Germany) were used in the study. The first two were research-grade devices while the third was the iThera MSOT Acuity Echo CE, approved for clinical use in the European Union and the United Kingdom. To minimise any potential differences in overall signal intensity due to slight differences in hardware, single-wavelength comparisons were made only between scans made on the CE device, while data from all devices were included in analysis of wavelength signal ratios.

**Figure 1.**
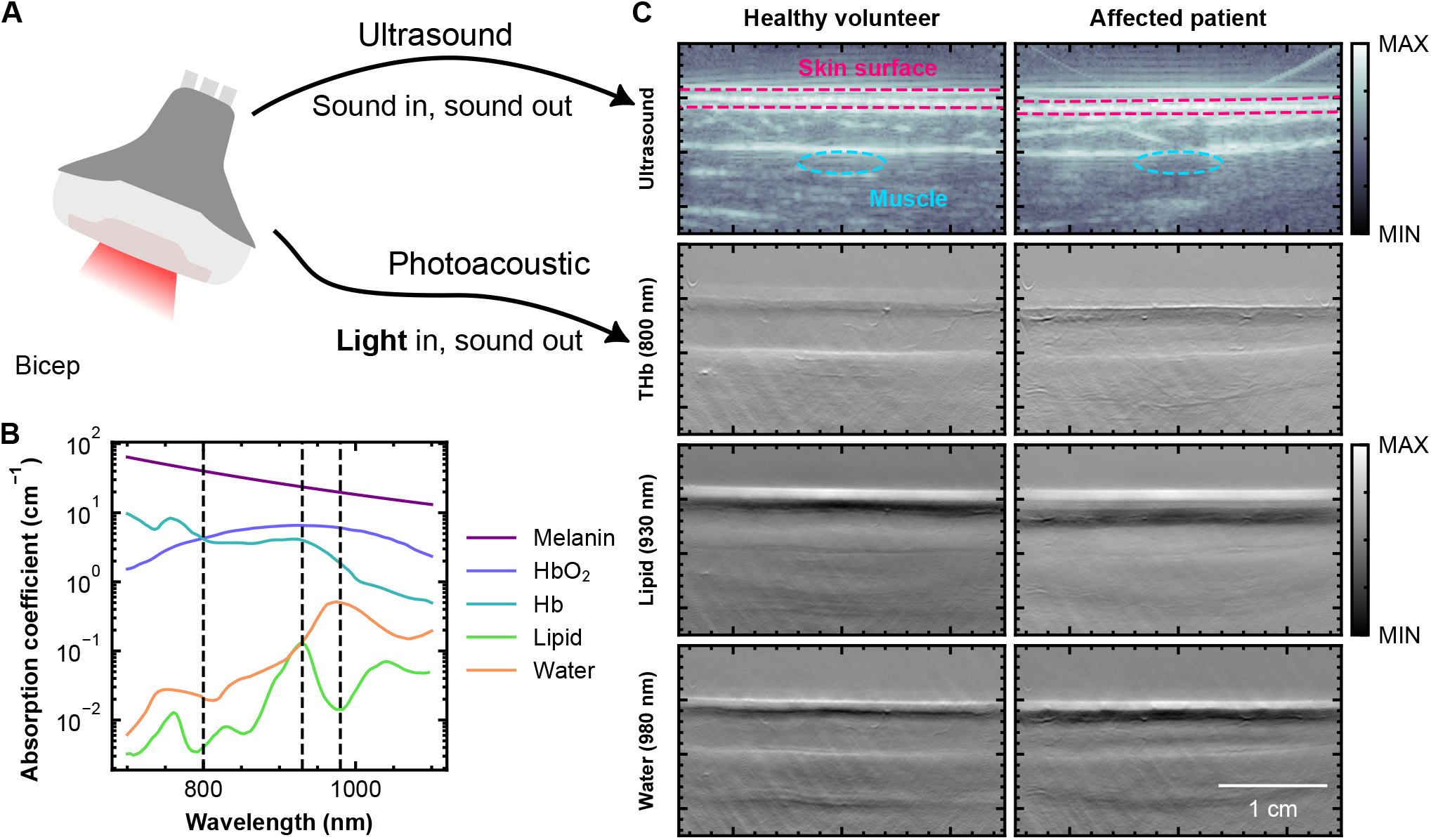
Study overview. **A:** The bicep of the study participants was measured using ultrasound and photoacoustic imaging modes in the same system. **B:** Wavelength-dependent optical absorption spectra of several key molecules targeted in this study, with wavelengths used in visualisations shown with a grey dotted line. **C:** Example ultrasound and photoacoustic images. Photoacoustic images are shown at wavelengths near the isosbestic point of haemoglobin (800 nm) and the peaks of lipid (930 nm) and water (980 nm) absorption.

Each imaging system had equivalent underlying hardware components and captured both ultrasound and PAI data, as extensively described in previous reports.^25^ Briefly, light excitation was provided by a neodymium-doped yttrium aluminium garnet (NdYAG) laser with a pulse repetition rate of 25 Hz. The laser output wavelength was tuned between 660 nm and 1300 nm to target the absorption spectra of different molecules. Laser light was delivered by a single optical fibre. The laser output energy was maintained below 30 mJ, as required by maximum permissible exposure limits. Acoustic waves were detected by an arc-shaped array of ultrasound transducers, covering a 120-degree angle. B-mode ultrasound images were acquired in parallel, giving co-registered anatomical information alongside the functional information from PAI.

Photoacoustic images were reconstructed in Python using the PATATO toolkit^26^ and custom-written code (https://github.com/tomelse/MITOX). A model-based image reconstruction algorithm was used to generate two-dimensional images, with a 4 cm field of view and pixel size of 100 μm. For quantitative analysis, B-mode ultrasound images were annotated with polygon regions of interest around the skin surface and ellipsoidal regions of interest of equal area (major axis 4 mm, minor axis 1 mm) were constructed at the top of the muscle. The top of the muscle was chosen as it is closest to the skin surface and therefore minimises the spectral colouring and signal intensity degradation associated with light attenuation with depth.

To investigate the photoacoustic signals associated with different molecular biomarkers, we tested photoacoustic signal intensities at wavelengths near absorption peaks and their ratios, and evaluated the outputs from linear spectral unmixing, where the multi-wavelength intensities of each pixel are expressed as a linear combination of the optical absorption spectra of oxy-haemoglobin (HbO_2_) and deoxy-haemoglobin (Hb).^27^ The output signals from PAI cannot yield absolute concentrations due to incomplete information regarding the light fluence in tissue, however, spectral unmixing, single-wavelength quantification and ratio quantification strategies have yielded promising biologically relevant biomarker data across a range of clinical studies.^14,28–30^ We evaluate both strategies here, using the mean photoacoustic signal from each acquired wavelength across the manually drawn regions of interest. Negatively valued pixels, a non-physical artefact of imperfect photoacoustic image reconstruction were excluded from our analyses.^31,32^

Due to the substantial influence of melanin absorption on photoacoustic image quality and quantitative measures, we evaluated the individual typology angle (ITA) at the imaging site for each participant. ITA was either measured using a colorimeter (Colorimeter DSM-4, Cortex Technology, Aalborg, Denmark) or interpolated from the photoacoustic signal at 700 nm averaged across the skin surface region of interest, as described previously.^21^

For linear spectral unmixing, the coefficients of each haemoglobin component, Hb and HbO_2_ were evaluated using the matrix pseudo-inverse method in the Python library NumPy and total haemoglobin was calculated as THb = HbO_2_ + Hb.^33^ We further estimated blood oxygenation as the ratio of oxyhaemoglobin to total haemoglobin (HbO_2_/THb). While this quantity correlates with blood oxygenation,^33–35^ as mentioned above, spatial variations in the light fluence mean that this is not strictly sO_2_. For this reason, we denote this quantity 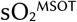.

### Statistical analysis

Statistical analyses were conducted in Python using the scientific Python library, SciPy. To avoid pseudo-replication, multiple values from the same patient were averaged before applying statistical tests. For all comparisons between two groups, e.g. between male and female groups, or healthy volunteers and m.3243A>G patients, the two-tailed permutation test was applied with 100,000 resamples (SciPy function `permutation_test`, with `statistic = mean(group 1) – mean(group 2)`) to allow for datasets that are not normally distributed. P-values smaller 0.05 were considered statistically significant. As this is an exploratory study, and many of the compared quantities are highly correlated, no multiplicity correction was applied to the p-values, except for the pairwise evaluation of different groups in our muscle weakness analysis. For pairwise comparisons between m.3243A>G patients with and without muscle weakness and healthy volunteers, we applied a conservative Bonferroni multiplicity correction using the Python library statsmodels.

## Results

### Correcting for the confounding effects of skin tone and sex enables quantitative assessment of PAI biomarkers

The high optical absorption of melanin in the skin, especially at short wavelengths (Figure 1B), can adversely affect quantification in PAI.^20–24^ We therefore ensured that the distribution of skin tones in healthy volunteers matched that of the m.3243A>G patients. Prior to matching the skin tone distribution between the healthy volunteers and m.3243A>G patients, the individual typology angle (ITA, a quantitative measure of skin tone, where higher is lighter) in the healthy volunteer group was significantly (p=0.01) lower than that of the affected patient group (mean ± standard deviation: healthy volunteers - 20.9° ± 35.8°, m.3243A>G patients - 48.0° ± 12.5°), suggesting that there was a much wider range of skin tones in the healthy volunteer cohort than the affected patient cohort. We excluded healthy volunteers with ITA lower than the lowest affected patient. Afterwards, there was no significant difference (p=0.844) between ITA values of the two groups (mean ± standard deviation: healthy volunteers - 48.9° ± 11.8°, m.3243A>G patients - 48.0° ± 12.5°). After the group refinement, there was also no significant difference between the two groups in the photoacoustic signal in the skin surface at any wavelength (p>0.121, Figure 2A and Supplementary Figure 1A).

**Figure 2.**
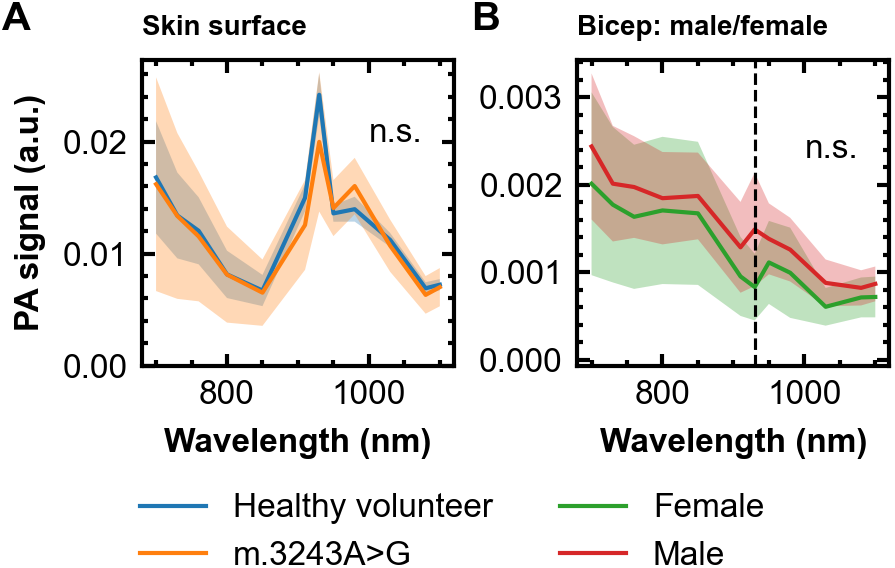
The potentially confounding effects of skin colour and sex were controlled for. **A:** Photoacoustic spectrum in the skin surface region of interest compared between healthy volunteers and m.3243A>G patients. **B:** Mean photoacoustic spectrum in the bicep muscle region of interest compared between male and female healthy volunteers. The shaded areas show two standard errors from the mean. The black vertical dotted line denotes the optical absorption peak of lipid (930 nm). “n.s.” denotes that there is no statistically significant difference between the groups at any wavelength.

The final dataset thus consisted of photoacoustic images from 21 healthy volunteers and 11 m.3243A>G patients. The two groups had a similar distribution of ages, skin colour (as determined by individual typology angle) and a variety of male and female participants (Table 1). In patients the mean value of heteroplasmy rate was 25% ± 18%; three patients were assessed as having muscle weakness, while eight did not (Table 1).

**Table 1:** Summary statistics for the study cohorts presented here after the exclusion of ineligible participants. Quantities are shown as mean ± standard deviation. Individual typology angle (ITA) is a quantitative measure of skin pigmentation, where higher corresponds to lighter skin.

|  | Healthy volunteers | m.3243A>G patients |
| --- | --- | --- |
| Number | 21 | 11 |
| Age (years) | 46 ± 19 | 43 ± 17 |
| Individual typology angle (°) | 49 ± 12 | 48 ± 13 |
| Sex: male / female | 8 / 13 | 5 / 6 |
| Heteroplasmy rate (%) | n/a | 25 ± 18 |
| Muscle weakness | n/a | 3 Yes, 8 No |

As the two study groups did not have the same number of male and female participants, we next explored the potential confounding effects of sex to reduce the chance of any incorrectly identified differences between patients and healthy volunteers. Using only data from the healthy volunteer cohort, the mean photoacoustic signals from the bicep did not show any statistically significant differences (p>0.151 for all wavelengths) between male and female healthy volunteers at any measured wavelength (Figure 2B and Supplementary Figure 1B). Nonetheless, while a clear lipid peak can be seen in the spectra of the male volunteers at 930 nm, a local minimum can be seen at the same wavelength in the female volunteers. Rather than an absence of lipid in the muscle in female patients, this spectral ‘dip’ is likely explained by a higher lipid concentration in the overlying tissue – the subcutaneous fat – leading to absorption of light at that wavelength before the light reaches the muscle.

### Single wavelength quantities and unmixed haemoglobin biomarkers do not vary significantly between healthy volunteers and patients

Having accounted for potential confounding effects, we first compared the raw intensity of the photoacoustic spectra at each wavelength in a region of interest defined at the top of the bicep muscle between healthy volunteers and patients (Figure 1C). No statistically significant differences (minimum p=0.237) were observed at any wavelength between the two groups (Figure 3A and Supplementary Figure 1C). Nonetheless, the photoacoustic spectra revealed interesting features that, while obscured by the high variance of the small dataset, prompted further investigation. For example, a trough at 930 nm was evident in the spectra of patients, consistent with lipid absorption and resembling the pattern observed in female healthy volunteers. Furthermore, a peak at 980 nm can be seen in the patients, corresponding to the optical absorption maximum of water (Figure 3A and Supplementary Figure 1C).

**Figure 3.**
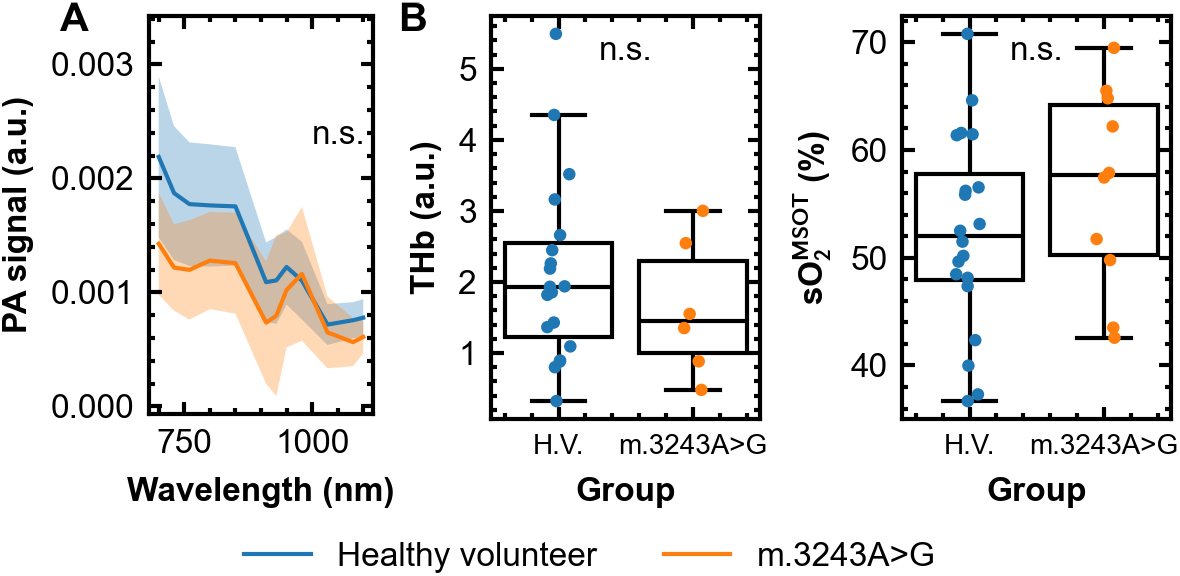
Single wavelength photoacoustic signal and unmixed quantities in the skin surface and bicep muscles of m.3243A>G patients and healthy volunteers. **A:** Mean photoacoustic spectrum in the bicep muscle compared between healthy volunteers and m.3243A>G patients. The shaded area shows two standard errors from the mean. **B:** Unmixed quantities, total haemoglobin (THb) and unmixed blood oxygenation 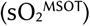 compared between healthy volunteers (H.V.) and m.3243A>G patients. “n.s.” denotes that there is no significant difference between the two groups shown; in panel A, there were no significant differences between signals at any wavelength.

Next, we estimated total haemoglobin (THb) and 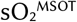 by linear spectral unmixing to assess blood content and oxygenation. These quantities were calculated pixel-wise and averaged across the region of interest at the top of the bicep muscle. No significant differences were observed in either of these quantities between healthy volunteers and m.3243A>G patients (p=0.42 for THb, p=0.24 for 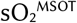, Figure 3B).

### Ratios between photoacoustic intensities differ significantly between healthy volunteers and patients and may be associated with muscle weakness

The small size of the patient cohort led to a high degree of variability in the raw signal intensities at each wavelength. To overcome the inter-subject variability, photoacoustic signal intensities from the bicep muscle were averaged at each measured wavelength and ratios were calculated pairwise between wavelengths. No substantial differences were observed between healthy volunteers and patients for pairs of shorter wavelengths (<930 nm) apart from 850/700 (p=0.031), however, many ratios between longer and shorter wavelengths did show significant differences (Figure 4).

**Figure 4.**
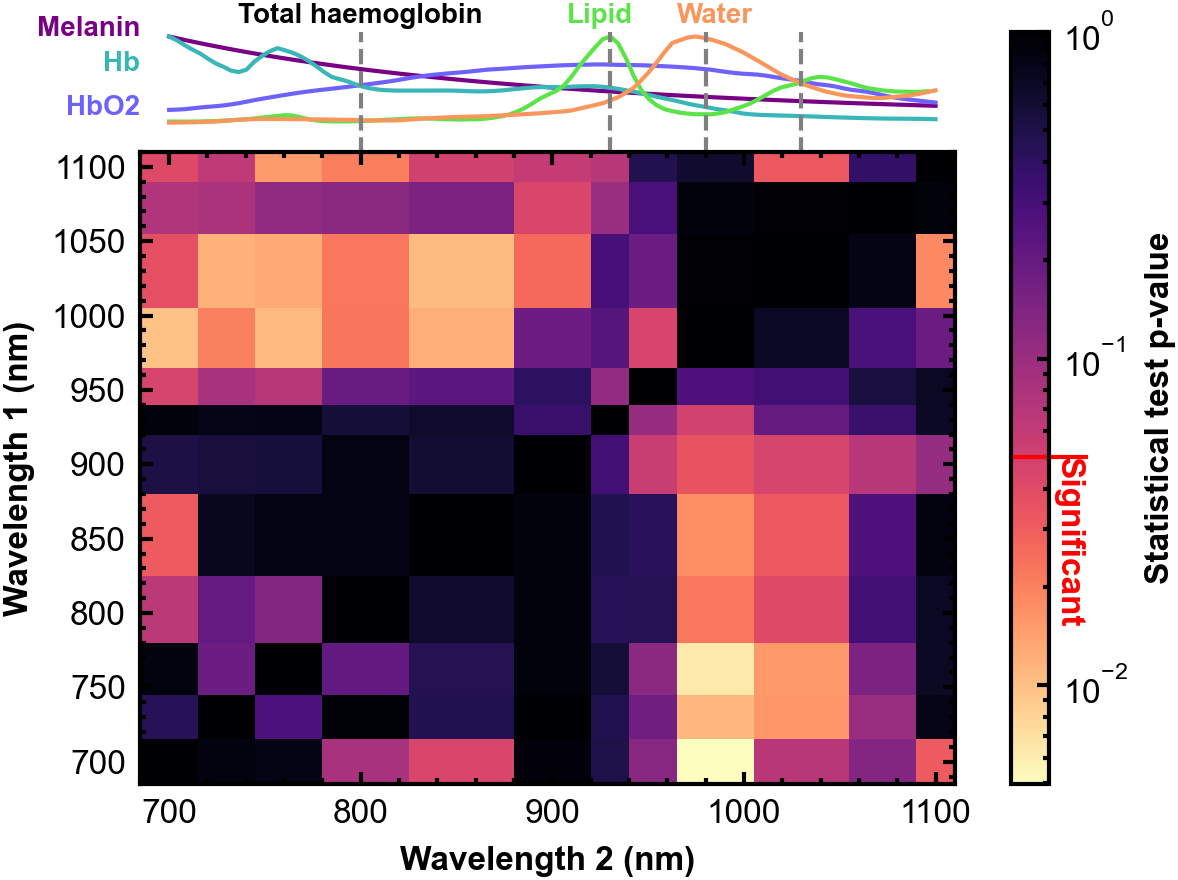
Ratios of photoacoustic intensities of the bicep muscle at certain wavelengths can differentiate between healthy volunteers and m.3243A>G patients. Heat map of the p-value from comparing the photoacoustic signal ratios at each pair of wavelengths between m.3243A>G patients and healthy volunteers. The normalised absorption spectra of the corresponding molecules are shown above the heat map on a linear scale. Hb: deoxy-haemoglobin, HbO_2_: oxy-haemoglobin.

Based on these findings, we chose ratios of key wavelengths that are associated with key biomolecules. We took two local maxima of lipid absorption (930 nm and 1030 nm) and a maximum of water absorption (980 nm) for more detailed comparison, taking the isosbestic point of oxy- and deoxy-haemoglobin (800 nm) as a normalisation wavelength (Figure 1A). The 1030/800 ratio showed a statistically significant difference between patients and healthy volunteers (p=0.021), as did 980/800 (p=0.025), and 930/980 (p=0.048), however 930/800 did not (p=0.58) (Figure 5A-D).

**Figure 5.**
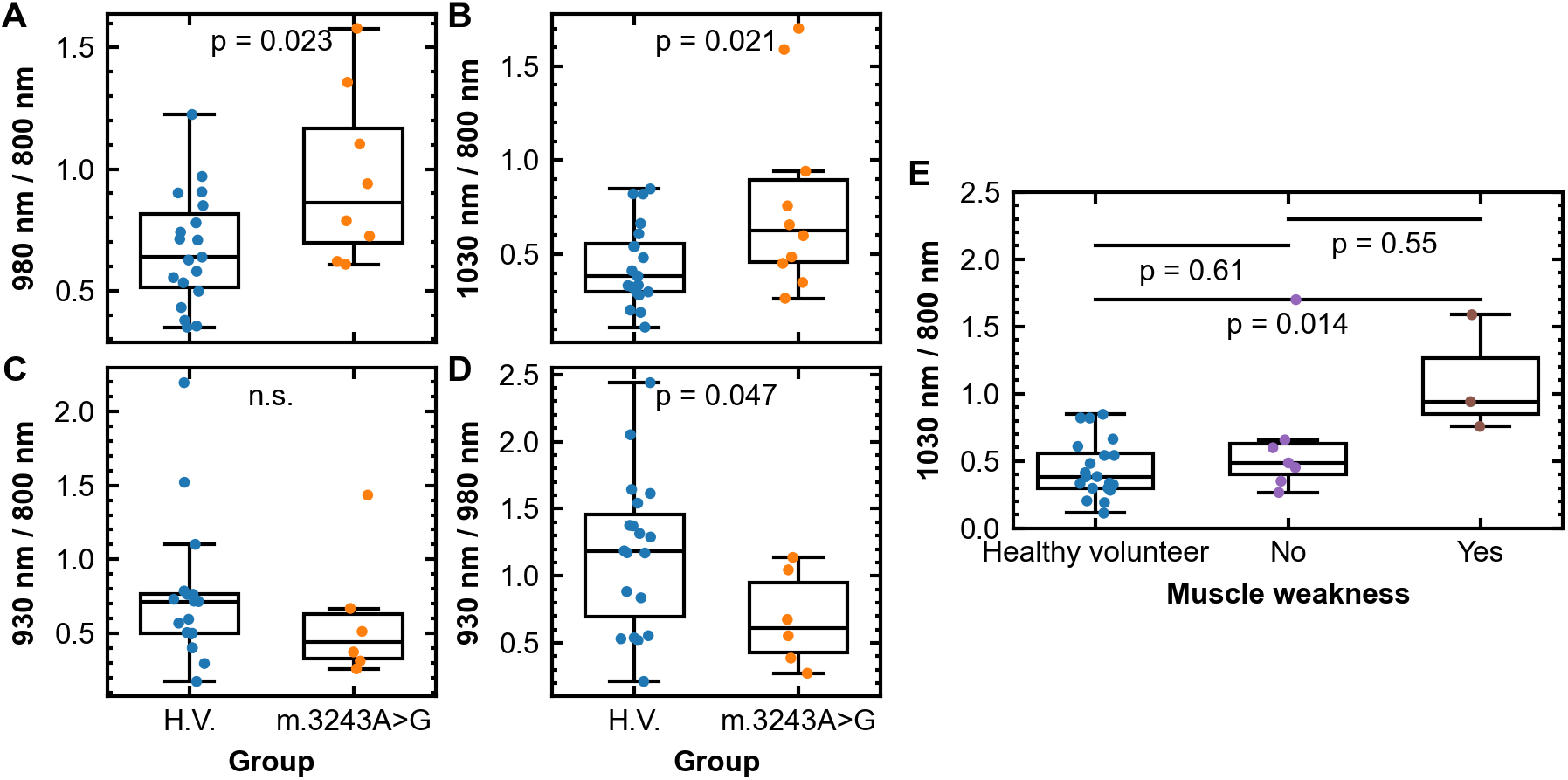
Ratios between pairs of singlewavelength photoacoustic signals in the bicep muscle differ between healthy volunteers (H.V.) and m.3243A>G patients. **A:** 980 nm (water peak) to 800 nm (total haemoglobin) ratio. **B:** 1030 nm (lipid local maximum) to 800 nm (total haemoglobin) ratio. **C:** 930 nm (lipid peak) to 800 nm (total haemoglobin) ratio. **D:** 930 nm (lipid peak) to 980 nm (lipid peak) ratio. **E:** 1030 nm peak to 800 nm peak ratio for healthy volunteers and m.3243A>G patients with and without muscle weakness. “n.s.” denotes that there is no significant difference between pairs of values (p>0.05). p-values in **E** are corrected for multiple comparisons by a conservative Bonferroni correction.

No statistically significant differences were observed between male and female healthy volunteers at any of the assessed wavelength pairs, nor were there significant differences in the same ratios calculated in the skin surface region, suggesting that changes were specifically observed in the bicep muscle (Supplementary Figure 2 and Supplementary Figure 3).

Finally, as a step towards using PAI to evaluate clinical severity, we evaluated the relationship between clinical parameters and the promising photoacoustic signal ratios. We compared the relationship between ratios of photoacoustic signal intensity between healthy volunteers and patients with and without muscle weakness. Interestingly, the 1030 / 800 ratio showed a significant increase (p=0.014) between the healthy volunteer group and the group with muscle weakness (Figure 5E).

## Discussion

The gold standard molecular biomarkers for mitochondrial disorders currently rely on invasive muscle biopsy, limiting our ability to monitor and treat these rare diseases. Existing non-invasive imaging methods rely largely on MRI or PET, which are expensive and their availability or applicability can be limited to monitor disease progression or treatment effect in clinical trials. As a non-invasive, contrast agent-free, low-cost imaging modality, we hypothesised that PAI could offer a convenient tool for molecular imaging of mitochondrial disease in skeletal muscle. We analysed PAI data from the bicep muscle of healthy volunteers and patients with a genetically defined mitochondrial disease (m.3243A>G) after controlling for potential confounding factors of skin tone and sex. Using these refined data, we evaluated differences in single-wavelength photoacoustic intensity in the bicep muscle, ratios between intensities at different wavelengths, and compared these to clinical observations.

Single wavelength values averaged across regions of interest in muscles have shown promise as a non-invasive biomarker of treatment response in other neuromuscular diseases, such as Duchenne muscular dystrophy, Pompe’s disease and spinal muscular atrophy.^14,17,18^ Here, we did not see any such difference between patients with mitochondrial disease and healthy volunteers, likely due to the small sample size and high variance observed in the dataset. Exploring ratios of different pairs of wavelengths allowed us to counteract the interpatient variability and revealed significant changes. Notably, we observe a significant increase in the ratio of the water peak (980 nm) wavelength to the isosbestic point of haemoglobin absorption (800 nm), and one of the lipid peaks (930 nm) and total haemoglobin (800 nm), between healthy volunteers and patients. The isosbestic haemoglobin peak in this case acts as a normalisation factor, as no significant difference was seen between the two groups in total haemoglobin from linear spectral unmixing.

We showed a relationship between a PAI ratio and clinical observations of muscle weakness. A statistically significant increase in the 1030 / 800 wavelength ratio was observed for m.3243A>G positive patients with muscle weakness, while no such increase was observed for patients without muscle weakness. 1030 nm corresponds to a peak of the lipid absorption spectrum, while 800 nm corresponds to the isosbestic point of oxy- and deoxy-haemoglobin, suggesting that this change may be associated with higher lipid levels in muscle tissue. This is in line with the histological detection of increased lipid droplets in ragged red fibres in mitochondrial myopathies.^36^ However, further validation would be required in a larger cohort to relate observed changes in imaging data to underlying differences in tissue composition.

While this study offers exciting paths for future research, the findings are preliminary and there are several key limitations to the study. Firstly, a practical limitation is that the number of patients is small, as is often the case for rare diseases. It would be valuable to expand the cohort to include longitudinal follow up data from these patients to enable stronger conclusions to be drawn. Secondly, direct correlations between photoacoustic intensity and molecular concentrations are challenging and an inherent technical limitation of PAI. In this study, the photoacoustic intensity in the region of interest was simply averaged at peaks of water and lipid absorption and the oxy- and deoxy-haemoglobin isosbestic point. It is well known that photoacoustic image intensity exhibits a non-linear relationship with the underlying molecule concentration, which depends on the optical properties of the overlying tissue. Future efforts to confirm these findings would therefore benefit from validation with a gold-standard technique, such as biopsy, or other established clinical imaging modality.

## Conclusion

Using PAI, we were able to identify several interesting biomarkers that could enable the non-invasive evaluation of mitochondrial disease status. Further study, however, is required to relate imaging biomarkers to underlying changes in tissue composition.

## Data Availability

All data produced in the present study are available upon reasonable request to the authors

## Acknowledgements

We would like to thank all patients and healthy volunteers for participating in this study. We also thank the CCRC nursing staff for their help with the study visits.

## Author contributions

Conceptualisation: TRE, SEB, RH

Formal analysis: TRE

Funding acquisition: RH, SEB, PFC

Investigation: TRE, LW, KS, MYT, CS, EA, CA, HB, EH, JvdA

Project administration: RH, PFC Software: TRE

Supervision: SEB, RH, PFC

Visualisation: TRE

Writing – original draft: TRE, SEB, RH

Writing – review and editing: JvdA

## Statements and declarations

### Ethical considerations

The study was conducted between May 2021 and October 2025, following approval by local Research Ethics Committees (Ref: 19/EE/0157; Ref: 13/YH/0310; Ref: 23/EE/0019). Written informed consent was obtained from all study participants. The study was conducted in accordance with the Declaration of Helsinki.

## Data availability

Data and code associated with this manuscript will be made available on the University of Cambridge data repository upon publication.

