## Supplementary Information for "Photoacoustic imaging in mitochondrial disease"

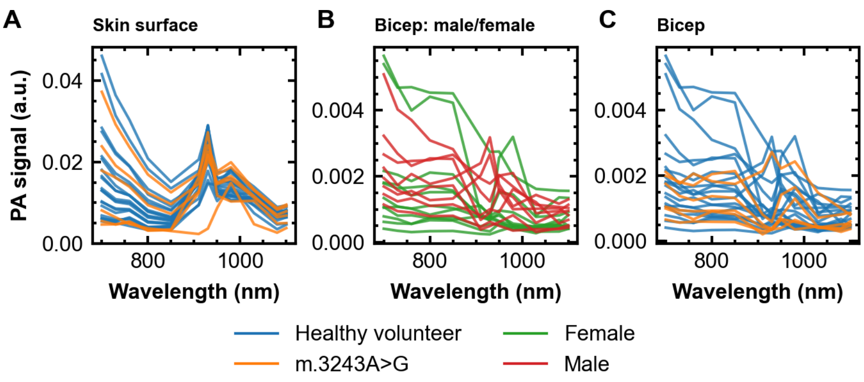


Supplementary Figure 1: **Plots of the photoacoustic spectra for everyone included in the study cohort. A:** Plots of the mean photoacoustic spectrum in the skin surface region of interest for each participant, grouped into healthy volunteers and m.3243A>G patients. **B:** Plots of the mean photoacoustic spectrum in the bicep muscle region of interest for each participant, grouped into female and male participants. **C:** Plots of the mean photoacoustic spectrum in the bicep muscle region of interest for each participant grouped into healthy volunteers and m.3243A>G patients.

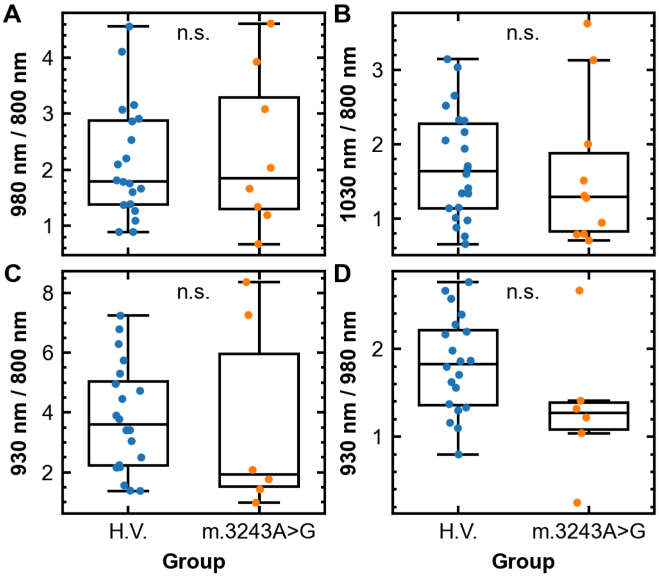


Supplementary Figure 2: **Comparison of ratios between healthy volunteers (H.V.) and m.3243A>G patients of photoacoustic signal values in the skin surface at all pairs of wavelengths. A:** 980 nm (water peak) to 800 nm (total haemoglobin) ratio. **B:** 1030 nm (lipid local maximum) to 800 nm (total haemoglobin) ratio. **C:** 930 nm (lipid peak) to 800 nm (total haemoglobin) ratio. **D:** 930 nm (lipid peak) to 980 nm (lipid peak) ratio. "n.s." denotes that there is no significant difference between pairs of values ($p>0.05$).

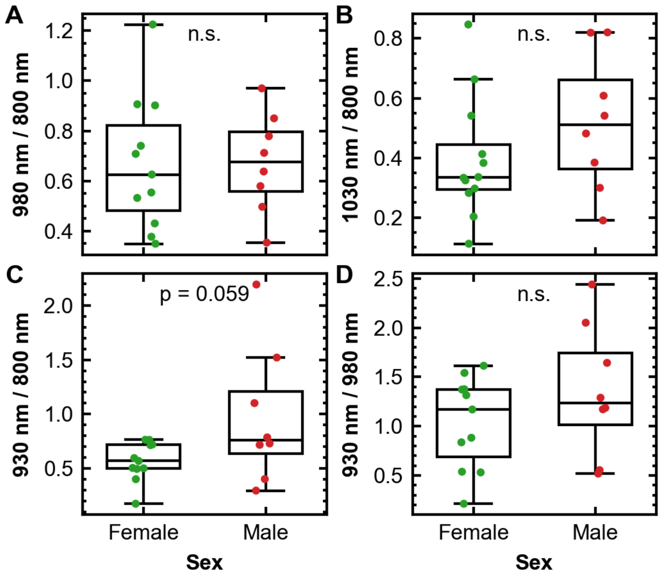


Supplementary Figure 3: **Comparison of ratios between male and female healthy volunteers of photoacoustic signal values in the bicep muscle at all pairs of wavelengths. A:** 980 nm (water peak) to 800 nm (total haemoglobin) ratio. **B:** 1030 nm (lipid local maximum) to 800 nm (total haemoglobin) ratio. **C:** 930 nm (lipid peak) to 800 nm (total haemoglobin) ratio. **D:** 930 nm (lipid peak) to 980 nm (lipid peak) ratio. "n.s." denotes that there is no significant difference between pairs of values ($p>0.05$).
